# The impact of COVID-19 control measures on social contacts and transmission in Kenyan informal settlements

**DOI:** 10.1101/2020.06.06.20122689

**Authors:** Matthew Quaife, Kevin van Zandvoort, Amy Gimma, Kashvi Shah, Nicky McCreesh, Kiesha Prem, Edwine Barasa, Daniel Mwanga, Beth Kangwana, Jessie Pinchoff, CMMID COVID-19 Working Group, W. John Edmunds, Christopher I Jarvis, Karen Austrian

**Affiliations:** Faculty of Public Health and Policy, London School of Hygiene and Tropical Medicine, London, UK; Faculty of Epidemiology and Population Health, London School of Hygiene and Tropical Medicine, London, UK; Health Economics Research Unit, KEMRI-Wellcome Trust Research Programme, Nairobi, Kenya; Centre for Tropical Medicine, Nuffield Department of Clinical Medicine, University of Oxford, Oxford, UK; Population Council, Nairobi, Kenya; Population Council, USA

**Keywords:** COVID-19, SARS-CoV2, social contacts, physical distancing

## Abstract

**Background:** Many low- and middle-income countries have implemented control measures against coronavirus disease 2019 (COVID-19). However, it is not clear to what extent these measures explain the low numbers of recorded COVID-19 cases and deaths in Africa. One of the main aims of control measures is to reduce respiratory pathogen transmission through direct contact with others. In this study we collect contact data from residents of informal settlements around Nairobi, Kenya to assess if control measures have changed contact patterns, and estimate the impact of changes on the basic reproduction number (*R_0_*).

**Methods:** We conducted a social contact survey with 213 residents of five informal settlements around Nairobi in early May 2020, four weeks after the Kenyan government introduced enhanced physical distancing measures and a curfew between 7pm and 5am. Respondents were asked to report all direct physical and non-physical contacts made the previous day, alongside a questionnaire asking about the social and economic impact of COVID-19 and control measures. We examined contact patterns by demographic factors, including socioeconomic status. We described the impact of COVID-19 and control measures on income and food security. We compared contact patterns during control measures to patterns from non-pandemic periods to estimate the change in *R_0_*.

**Findings:** We estimate that control measures reduced physical and non-physical contacts, reducing the *R_0_* from around 2.6 to between 0.5 and 0.7, depending on the pre-COVID-19 comparison matrix used. Masks were worn by at least one person in 92% of contacts. Respondents in the poorest socioeconomic quintile reported 1.5 times more contacts than those in the richest. 86% of respondents reported a total or partial loss of income due to COVID-19, and 74% reported eating less or skipping meals due to having too little money for food.

**Interpretation:** COVID-19 control measures have had a large impact on direct contacts and therefore transmission, but have also caused considerable economic and food insecurity. Reductions in *R_0_* are consistent with the linear epidemic growth in Kenya and other sub-Saharan African countries that implemented similar, early control measures. However, negative and inequitable impacts on economic and food security may mean control measures are not sustainable in the longer term.

**Research in context:** *Evidence before this study:* We conducted a PubMed search on 6 June 2020 with no language restrictions for studies published since inception, using the search terms (“social mix*” OR “social cont*” OR “contact pattern*) AND (“covid*”). The search yielded 53 articles, two of which reported changes in social contacts after COVID-19 control measures. The first study reported changes in contact patterns in Wuhan and Shanghai, and the second changes in contact patterns in the UK. We found no studies examining changes in contact patterns due to control measures in sub-Saharan Africa, and no studies disaggregating contacts by socioeconomic status.

*Added value of this study:* This is the first study to estimate the reproduction number of COVID-19 under control measures in sub-Saharan Africa using primary contact data. This study also moves beyond existing work to i) measure contacts in densely populated informal settlements, ii) explore how social contacts vary across socioeconomic status, and iii) assess the impact of control measures on economic and food security in these areas.

*Implications of all the evidence:* COVID-19 control measures have substantially reduced social contacts and disease transmission. People of lower socioeconomic status face greater transmission risk as they report more contacts. Control measures have led to considerable economic and food insecurity, and may not be sustainable in the long term without efforts to reduce the burden of control measures on households.

## Introduction

Over 6.7 million cases and 394,000 deaths from COVID-19 have been recorded worldwide as of 6 June 2020[1]. Most recorded cases and deaths have occurred in high-income countries in Europe and North America. Many countries introduced extreme physical distancing control measures to control SARS-CoV2 transmission[2]. Modelling studies suggest that without substantial mitigation measures, most low- and middle-income (LMIC) settings, including sub-Saharan Africa, will experience a delayed, but severe epidemic[3, 4]. Yet to-date, the numbers of recorded cases and deaths in Africa are much lower than predictions, prompting speculation on why many African countries have so far avoided a severe uncontrolled epidemic. A range of reasons has been proposed, including differences between settings in case and death detection capacity, demographic factors such as population age distribution, and the role of temperature and aridity in transmission[5–10]. However, many sub-Saharan African countries implemented lockdown and curfew measures far earlier in their country’s epidemic trajectories than most higher-income settings in Europe and North America. For example, Kenya – the focus of the current study – implemented a partial lockdown on 06 April 2020 when the country had recorded just 158 cases and 6 deaths. In contrast, although case detection rates may differ between settings, the UK implemented its own lockdown on 23 March 2020 after recording 6,650 cases and 335 deaths[1, 2]. The first reported case in Kenya was on 13 March 2020, and schools closed on 15 March 2020. Suspension of international flights, including mandatory quarantine of incoming residents, closure of bars and restrictions on restaurant opening hours, and a ban on large gatherings were imposed on 25 March 2020, soon followed by an enactment of a nationwide curfew from 7pm to 5am. On 5 April 2020, the Kenyan government declared wearing face masks as mandatory in any public place. Recently, cessation of movement was imposed in informal settlements in Mombasa and Nairobi, following a rise in cases in Nairobi’s Kibera informal settlement. Consequently, the government has indicated additional physical distancing measures may be authorised.

Physical distancing control measures seek to reduce the number of contacts between people where transmission could occur. To predict the impact of control measures accurately, quantitative data on the number and type of contacts between people is required. To-date, only a few empirical studies have been published to assess the impact of COVID-19 control measures on contacts; these have been conducted in China[11], the USA [12] and Europe [13]; but none were undertaken in sub-Saharan Africa. In fact, prior to the current pandemic, a systematic review[14] reported that just four social contact surveys out of 64 had been conducted in sub-Saharan Africa, including one in Kenya[15–17]. To our knowledge, just one LMIC study has been published since this review[18]. This lack of evidence means that many SARS-CoV-2 transmission models primarily use synthetic contact matrices for LMIC settings, which use demographic, household composition, classroom size and other data to adjust social contact data from primarily high-income settings[19, 20]. Although one social mixing study was conducted in Kilifi, a coastal area of Kenya[21], outside of one study which collected data from a South African township[16], no published contact data exist from informal settlements, which may be particularly vulnerable to COVID-19 due to high levels of population density, indoor crowding and household sizes, alongside intergenerational mixing within the household.

Between-person contacts drive the transmission of respiratory pathogens, such as SARS-CoV-2. Understanding how contact patterns change under different control measures is important to inform decisions on whether and how to implement them. In this study, we describe a survey of contact patterns conducted among a sample of adults from five informal settlements in urban and peri-urban areas around Nairobi. We explore how direct contacts vary across respondent characteristics, including by socioeconomic status. We estimate the impact of current control measures on the reproduction number, *R_0_*, to evaluate whether these measures might be sufficient to control the epidemic. We also describe income losses and food security that respondents attribute to COVID-19 and control measures.

## Methods

### Ethics

Participation in the study was voluntary and analyses were conducted on anonymised data. The study was approved by the internal review board of the Population Council (study number 936), the ethics committee of the London School of Hygiene and Tropical Medicine (reference number 22294), and the AMREF Health Africa Ethics and Scientific Review Committee in Kenya (P803/2020).

### Survey methodology

Adult respondents were recruited from two existing Population Council cohorts in five informal settlements around Nairobi (Kibera, Huruma, Kariobangi, Dandora, and Mathare). The existing cohorts were part of the Adolescent Girls Initiative Kenya (AGI-K) and Nisikilize Tujengane (NISITU – Listen to Me, Let’s Grow Together) studies. The cohorts were in place to study the impacts of multi-sectoral interventions on adolescents, and consisted of randomly-selected households from informal settlements which contained at least one adolescent in January 2015 (AGI-K) or January 2018 (NISITU). In May 2020, 1750 respondents from AGI-K and NISITU cohorts completed a telephone survey on COVID-19 knowledge, attitudes, and perceptions (KAP). Of these 1750, an age and sex stratified random sample of 213 respondents completed a contact survey. Stratification was based on 2019 Kenya census data for Nairobi county, with a target sample size of 200 and 20% oversampling to account for refusal. This was based on the sample sizes of similar contact surveys[14], alongside feasibility of phone interviewing during lockdown. Background data, including household ownership of assets, were merged from previous survey rounds. Respondents were first asked a range of questions on COVID-19 including knowledge and experience of testing and symptoms, economic impacts on the household and food availability and cost. Then respondents were asked to report all direct physical and non-physical contacts made between 5am the day preceding the survey and 5am the day of the survey. A direct contact was defined as someone respondents met in person and with whom they had either i) “*physical contact (any sort of skin-to-skin contact e.g. a handshake, embracing, kissing, sleeping on the same bed/mat/blanket, sharing a meal together out of the same bowl, playing football or other contact sports, sitting next to someone while touching shoulder to shoulder, etc*.”, or ii) “*Non-physical contact (you did not touch the person, but exchanged at least a few words, face-to-face within 2 metres – for example, someone you bought something from in the market, or rode with on a minibus, or worked with in the same area)*”. All respondents were over the age of 18 so no contact data were collected from children, however respondents were able to list contacts under the age of 18.

We made pragmatic adaptations to existing contact measurement tools to allow them to be conducted over the phone, primarily to reduce respondent burden and to ensure that aggregate contact data were not biased downwards by respondent fatigue. Respondents were first asked about contacts with members of their household the previous day, recording the contact age, gender, and whether contacts were physical or non-physical. Then respondents were asked how many non-household contacts they had had in the same timeframe. Those who reported nine or fewer outside household contacts were asked to describe each contact’s age, gender, whether the contact was physical or non-physical, the duration of the contact, and whether a mask was worn by the respondent or contact. Those who reported ten or more outside-household contacts were asked how many of these contacts were physical/non-physical, in the age ranges under 18, 18–60, and over 60. The contact tool is shown in supplementary file 1.

### Statistical analysis

R version 4.0.0 and Stata 15 were used for analyses; the code and data are publicly available here. The age and gender of respondents were compared to the full sample from which they were drawn, alongside census data to assess the representativeness of the sample. Data on household assets were used to classify respondents into wealth quintiles using principal component analysis; supplementary file 2 gives information on this, alongside methods used to estimate economic and food security.

We calculated the mean number of social contacts per person per day, stratified by respondent age, sex, household size, and education level. We then calculated social contact matrices for the age category-specific daily frequency of direct contacts, adjusting for contact reciprocity and the age distribution using census data from informal settlement sub counties. We then compared the mean total number of daily contacts by age group to the only empirical dataset available from Kenya in Kiti et al.[21], alongside synthetic matrices from 2017 [19] and 2020 [20]. Kiti et al. collected data on physical contacts only, so we restrict our sample to physical contacts when comparing with this study. We adjusted both matrices to match the age structure of the informal settlement setting, using the 2019 Kenyan Population and Housing Census to adjust from Kilifi and nationally representative populations respectively[22] supplementary file 3 provides more detail. Because Kiti et al. collected data on the age of contacts in categories (< 1, 1–5, 6–15, 16–19, 20–49, 50+) which were different to those in this survey, we restructured both age matrices and used 1000 bootstrapped samples of both datasets to impute the number of contacts for matching age ranges. We adjusted for symmetry after bootstrapping because one age range in our data (60+) had fewer than five respondents. Bootstrapping was not possible with Prem et al. matrices as they do not relate to individual level data.

As respondents under the age of 18 were not included as survey respondents, we imputed child contacts using methods developed by Klepac et al.[23], and implemented for the same purpose in a UK study[13]. This involved taking the ratio of the dominant eigenvalues between our matrices and the comparable setting-adjusted matrices to scale missing matrix elements.

Finally, we estimated the impact of control measures on the basic reproduction number (*R_0_*) in this population. Because there are no baseline contact data from this population without control measures, we assume that contact patterns in this sample prior to control measures were similar to those estimated by Kiti et al. or Prem et al. We make the common assumption for respiratory infections that the next generation matrix is a function of the age-specific number of contacts, the per-contact transmission probability, and the duration of infectiousness, and that *R_0_* is therefore proportional to the dominant eigenvalue of the contact matrix[18][17].We assume that existing matrices are comparable to the informal settlement setting of this study, that there were no changes in the duration of infectiousness during the study period, that per-contact transmission probability also remained constant, and that all age groups have the same per contact transmission probability, given infection. With these assumptions, the relative reduction in *R_0_* can be estimated as the reduction in the dominant eigenvalue of the contact matrices. Finally, our central estimate of the *R_0_* of SARS-CoV2 is 2.6 (SD = 0.54), as estimated in a meta-analysis of published estimates of *R_0_* prior to the introduction of control measures[13]. Because studies in this meta-analysis were predominantly based on European and Asian countries, we explore a lower bound of 1.46 (SD = 0.38) based on the earliest estimate of the time-varying reproduction number in Kenya[24].

## Results

### Respondent and contact characteristics

Out of the 1970 people sampled for the KAP survey, 1745 interviews were completed. Of the initial 1970 sampled, 237 were sampled to complete the additional contacts module. In total, 213 were successfully interviewed and recorded 3809 contacts. 830 (22%) of these were household contacts, and 324 (9%) were non-household contacts on which we have detailed information. The remaining 2655 (70%) were non-household contacts of respondents who reported ten or more such contacts. The mean age of respondents was 33 (SD: 11.38, max: 70) and 51% were female (108/213). Table 1 shows that the age and gender distribution of respondents broadly matched that of a) the sample from which respondents were randomly chosen, and b) the Kenyan adult population. Compared to both groups there is some indication that our sample has more 18–29 year olds and fewer 60+ year olds than national data, whilst our sample is substantially older than that of Kiti et al.

**Table 1:** Respondent characteristics in this study and comparison with data from mixing module respondents, full sample, and Kenya national demographics

|  | Respondents in this survey (n=213) |  | KAP survey respondents (n=1745) | Kenya* (all ages) | Kenya* (>19 only) | Kiti et al. 2017 |  |
| --- | --- | --- | --- | --- | --- | --- | --- |
| **Age group** |  |  |  |  |  | **Age group** |  |
| 0-17 | 0 | - | - | 50%* |  | <1 | 15% |
| 18-29 | 95 | 44% | 28% | 18%* | 36%* | 1-5 | 16% |
| 30-39 | 61 | 29% | 30% | 14% | 28% | 6-14 | 17% |
| 40-49 | 34 | 16% | 29% | 9% | 18% | 15-19 | 16% |
| 50-59 | 21 | 10% | 10% | 5% | 10% | 20-49 | 24% |
| 60+ | 3 | 1% | 3% | 4% | 8% | 50+ | 11% |
| **Gender** |  |  |  |  |  |  |  |
| Male | 106 | 50% | 37% | 50% | 51% | Male | 46% |
| Female | 108 | 50% | 63% | 50% | 49% | Female | 54% |

\*Kenyan national data from United Nations World Population Prospects [19]

### Implications of COVID-19 control measures

Eight respondents (4%) reported two or more COVID-19 symptoms^1^ in the previous seven days. 42% of respondents (89/213) thought they had a high chance of acquiring SARS-CoV2, and 81% (172/213) thought the implications would be “severe” or “very severe” if they caught the virus. When asked an open ended question without prompting what they would do if they developed COVID-19 symptoms, 64% (136/213) thought they would take a test, and 7% (16/213) said they would stay at home or avoid social gatherings. Just 6% (13/213) of respondents knew someone who was either suspected of having COVID-19 or who had tested positive.

Respondents reported substantial food and economic insecurity due to COVID-19 and control measures. Around a third (36%, 76/213) reported the pandemic had caused a complete loss of income, and an additional 50% (107/213) reported partial income losses. 83% (177) reported experiencing increases in food prices, and three-quarters of respondents reported eating less or skipping meals due to having too little money for food (74%, 158/213); all but one (157/158) reported that this was due to the situation with COVID-19. Just 21% (44/213) reported receiving monetary or non-monetary assistance in the previous seven days – 78% (166) reported that food was the one of the biggest needs that was currently unmet.

COVID-19 control measures meant 92% (196/213) of respondents reported seeing friends less, and 64% (136) seeing family less. 25% of respondents (54/213) reported leaving the settlement where the interview was conducted in the previous 24 hours. At the time of data collection, mask wearing was required by the Kenyan government in public places and was very common: 94% (199/211) of respondents reported “always” wearing a mask outside of their house.

### Contact patterns

The mean number of contacts reported was 18 (median 13, IQR 7–23), 4 household contacts (median 4, IQR 3–12) and 15 non-household contacts (median 10, IQR 4–20). As shown in figure 1, respondents in the poorest quintile reported 1.5 times as many contacts as those in the richest quintile and we find evidence of a downwards trend in contacts as socioeconomic status increases (non-parametric test for trend p = 0.02). There was weak evidence that men had more contacts than women (20.3 – 15.5 = 4.8, t-test p = 0.04) and contacts increased with age (non-parametric test for trend p = 0.05). Just 22% (847/3841) of contacts were reported within the household, and total contacts did not vary substantially by household size or by respondent education level. This lack of variation by household size is consistent with most contacts being outside of the household.

**Figure 1:**
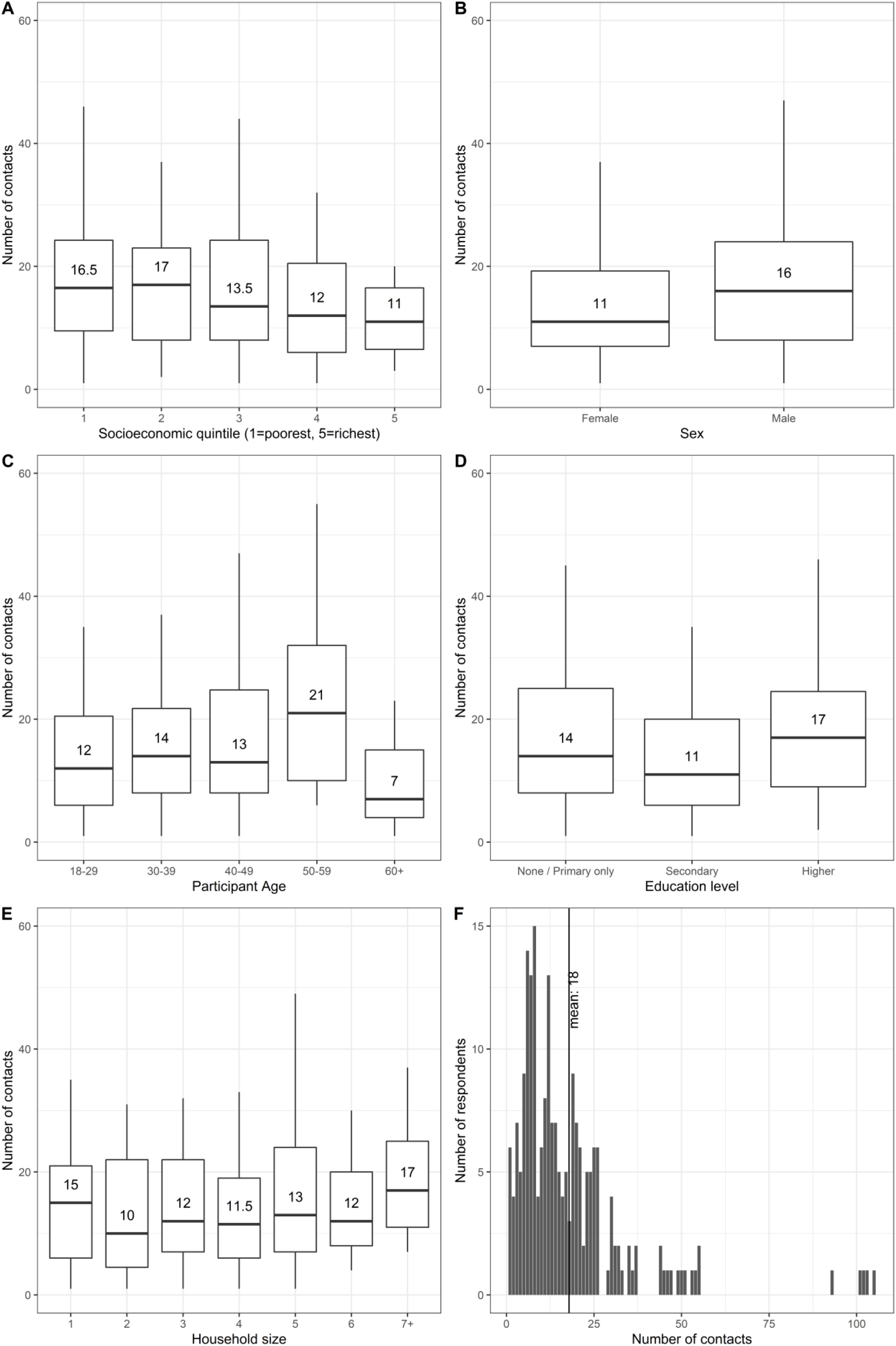
Median number of direct contacts (physical and non-physical) by (A) socioeconomic status quintile, (B) gender, (C) respondent age, (D) education level, and (E) household size. Each panel shows the median, hinges (25th and 75th percentiles), and whiskers representing upper and lower adjacents. Outliers are not displayed in boxplots for scale, these are plotted in (F) showing the distribution of the number of direct contacts reported.

Figure 2 summarises the characteristics of contacts for which we have detailed information (830 household contacts and 324 non-household contacts where a respondent reported fewer than ten non-household contacts). Most physical contacts were household contacts and the proportion of female contacts was higher among household than non-household contacts. Just 8% (27/324) of all contacts took place without a mask being worn by either the respondent or the contact. Most reported contacts were brief: 40% (130/324) were under five minutes, and a further 23% (75/324) between five and 15 minutes. Finally, 41% (133/324) of non-household contacts took place in an outside location, and 34% (110/324) of non-household contacts were in the home of the respondent or contact. Figure 3 shows age-specific contact matrices disaggregated by contact location and type; these are asymmetric and not adjusted for demography. Matrices are consistent with the majority of contacts occurring outside of the household and being non-physical.

**Figure 2:**
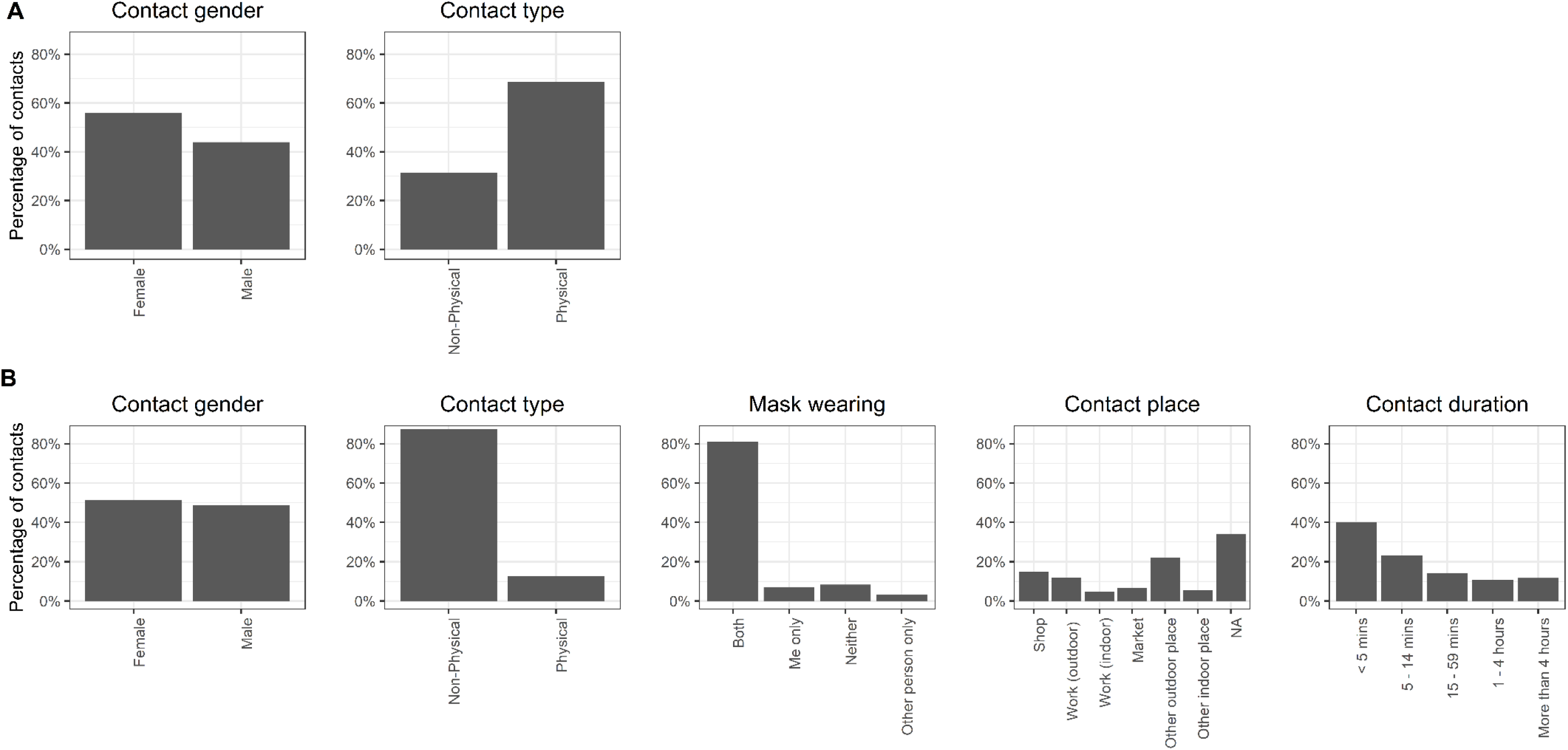
Characteristics of (A) household and (B) non-household contacts for which full information was gathered

**Figure 3:**
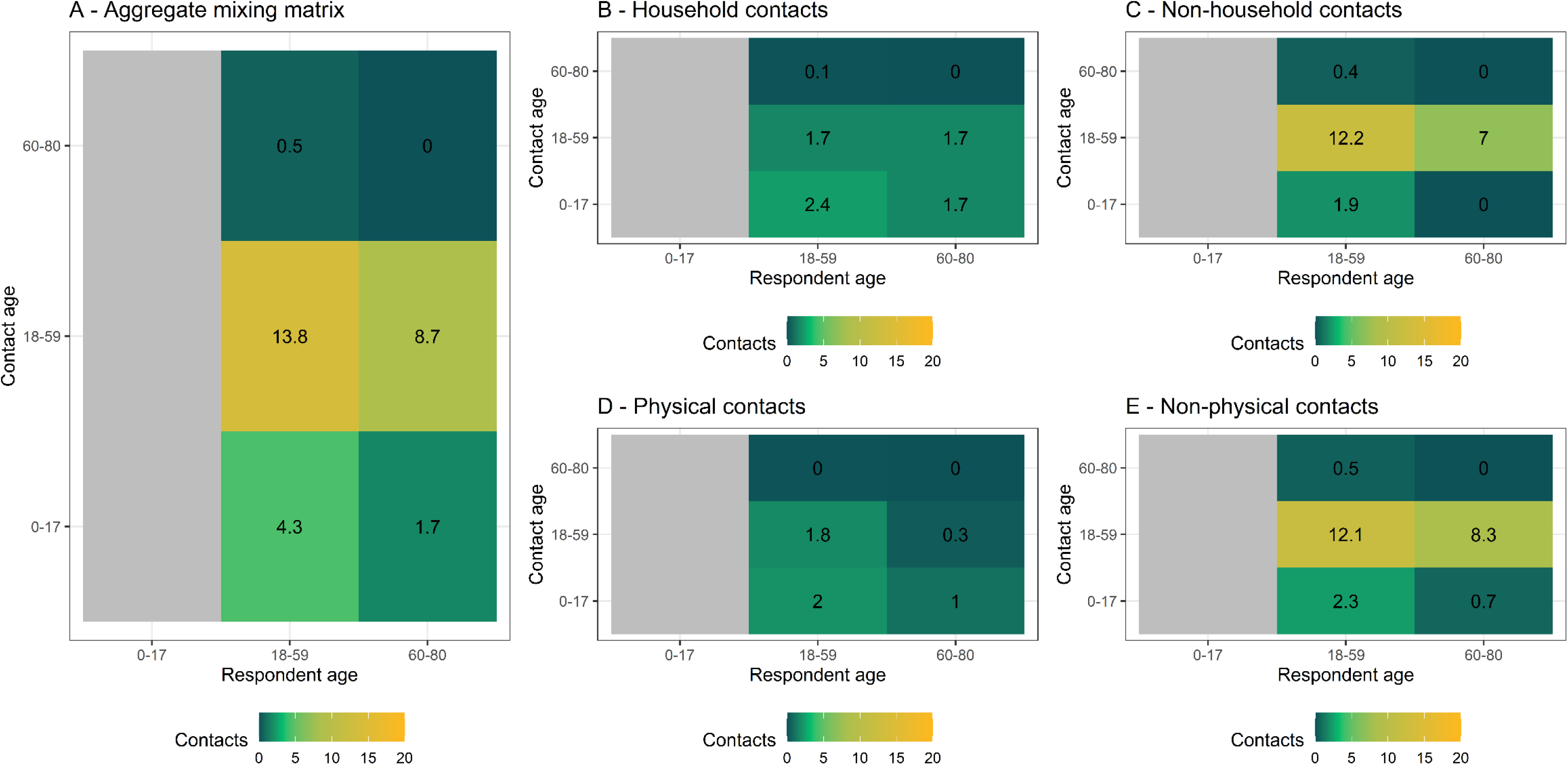
Age-stratified mean number of reported contacts from survey respondents recruited from five informal settlements around Nairobi. (A) is the aggregate mixing matrix, (B) shows household contacts only, (C) shows non-household contacts only, (D) shows physical contacts only, and (E) shows non-physical contacts only

Figure 4 uses the two existing contact matrices for Kenya to impute contact patterns for under 18s, adjusting for age-distribution and symmetry. The two pre-COVID-19 data sources differ substantially in their methods, and the differences are propagated in these adjusted matrices. We find a 62% reduction in physical contacts, and a 63–67% reduction in all contacts compared to before the epidemic. We estimate *R_0_* under control measures, shown in figure 5. All comparisons to pre-COVID-19 matrices assuming *R_0_* = 2.64 suggest that control measures reduced *R_0_* to below one, to 0.6 (IQR: 0.50, 0.68) for physical contacts and to either 0.54 (IQR: 0.46, 0.61) or 0.67 (IQR: 0.57, 0.76) depending the synthetic matrix used as comparator, based on Prem et al. 2017 [19] and 2020 [20] respectively. Using the lower R_0_ estimate of 1.46, we estimate reductions to 0.33 (IQR 0.27, 0.39) for physical contacts, and either 0.30 (IQR: 0.24, 0.35) or 0.37 (IQR: 0.3, 0.43) all contacts. Based on these values, control measures would have reduced the mean estimate of *R_0_* to below one even if the initial *R_0_* had been as high as 4.36 assuming only physical contacts lead to transmission, or either 3.9 or 4.8 assuming all contacts are equally risky.

**Figure 4:**
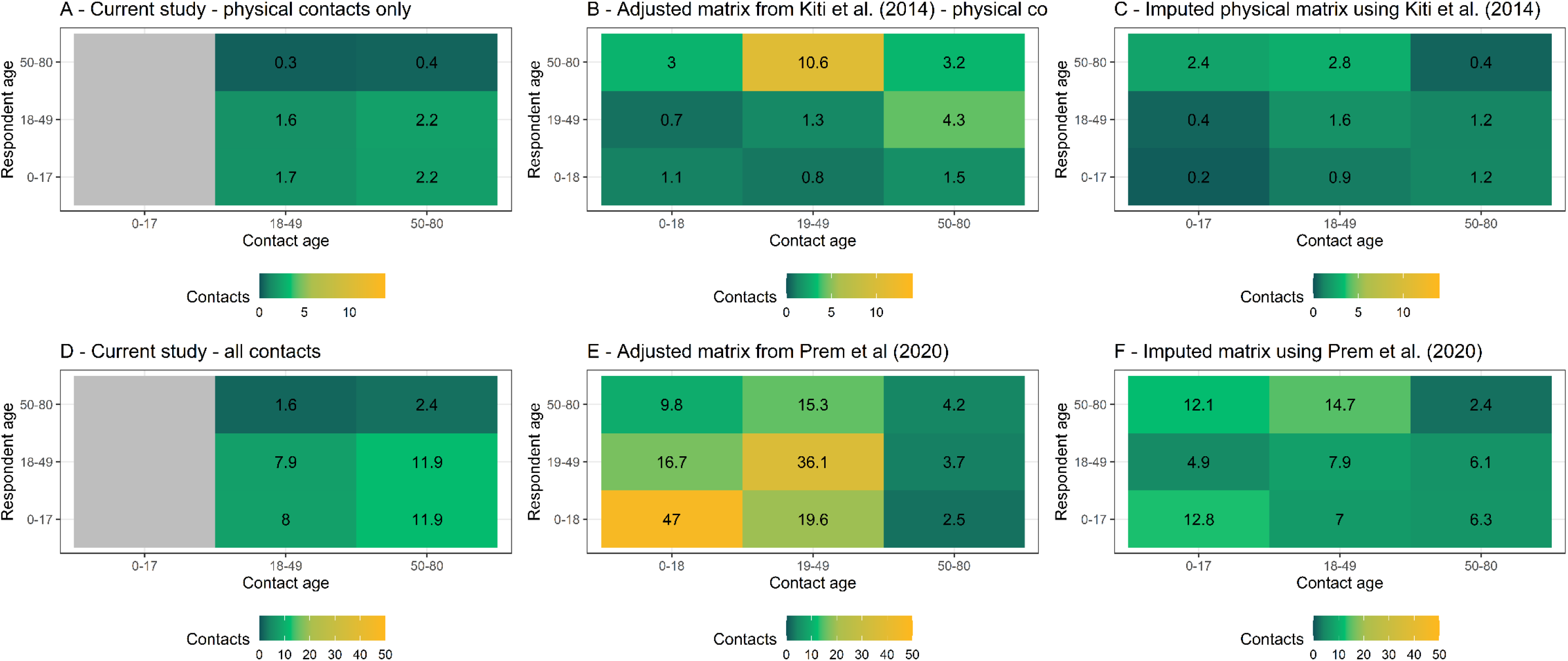
Mixing matrices with 1000 bootstrapped samples, where (A) is the unadjusted physical contact matrix, (B) the physical contact matrix from Kiti et al. (2014) adjusted for the age distribution of the informal settlement setting, and (C) is the mixing matrix produced when Kiti et al. data are used to impute child contacts. (D) is the unadjusted contact matrix, (E) the contact matrix of Prem et al. (2020) adjusted for the age distribution of the informal settlement setting, and (F) the mixing matrix produced when Prem et al. data are used to impute child contacts

**Figure 5:**
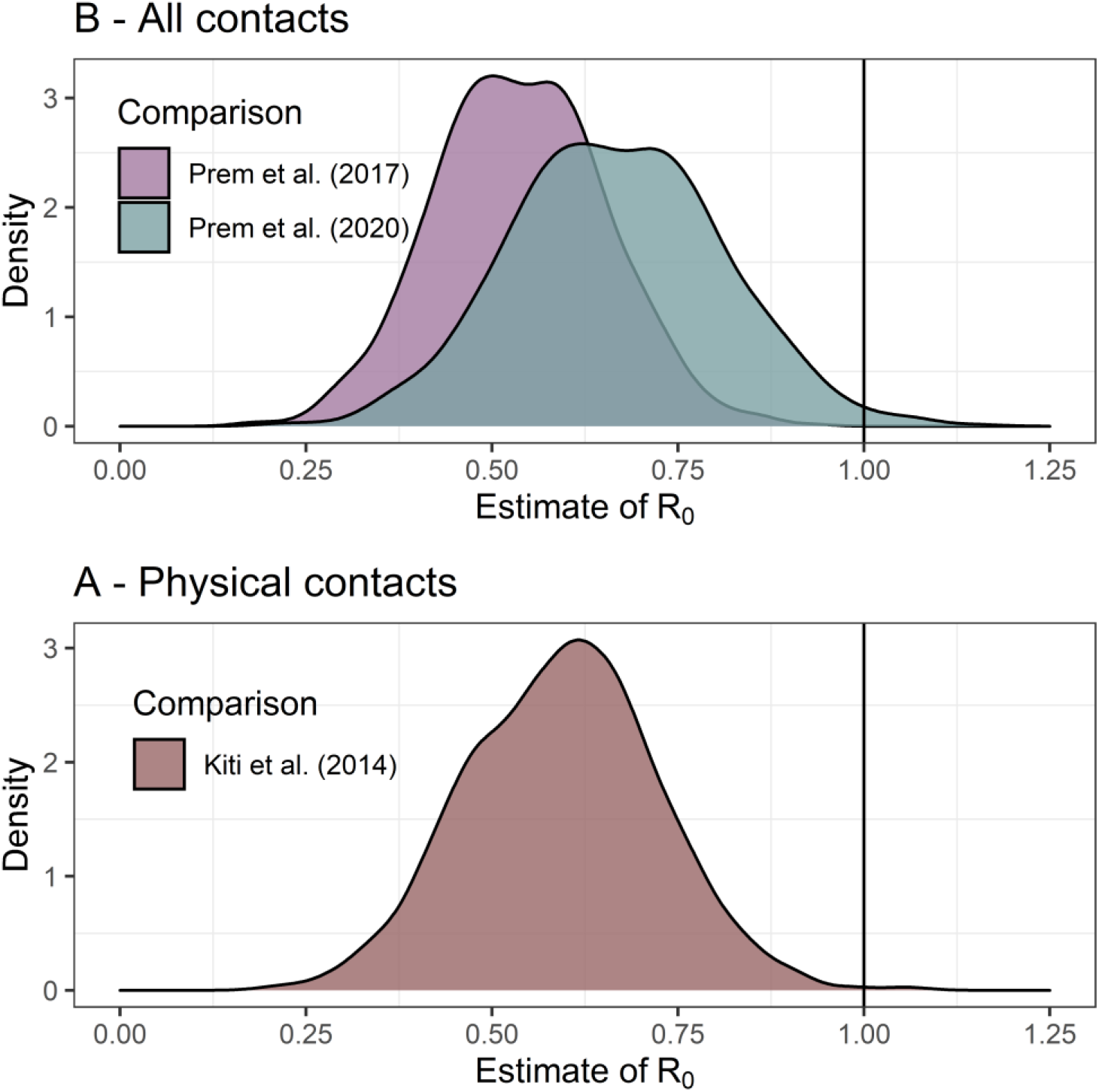
Estimated value of *R_0_* at time of survey Nb. *R*_0_ assumed ~Norm (2.6, SD = 0.54) prior to control measures

## Discussion

COVID-19 control measures in informal settlements appear to have led to a large reduction in social contacts. We find a 62–67% reduction in eigenvalues of contact matrices depending on the pre-COVID-19 matrix used; assuming an *R_0_* of 2.6, this would translate to an *R_0_* of between 0.5 and 0.7 at the time of data collection. By contrast, simulation estimates of the *R_0_* in an unmitigated COVID-19 epidemic in Kenya were between 1.78 (95%CI: 1.44 – 2.14) and 3.46 (95% CI: 2.81–4.17)[25]. The *R_0_* we estimate here is consistent with the linear shape of the Kenyan epidemic to-date. The large reductions in contacts we estimate are of similar magnitude to those seen in both the UK[13] – (74% reduction in contacts), in Wuhan and Shanghai[11] (86% reduction), and the USA (70% reduction) [12]. We are not aware of any comparable post-lockdown studies from low- or middle-income settings to-date, including sub-Saharan Africa.

Considerable food and economic vulnerability was reported due to COVID-19 control measures. Over 80% of respondents reported a partial or complete loss of income, and three quarters reported eating less or skipping meals due to COVID-19. Households reported they were receiving some assistance, but that their biggest remaining unmet need was food. Although the prevalence of COVID-19 was low, and these factors can largely be attributed to control measures rather than illness from COVID-19 itself, it is important to recognise the counterfactual of no control measures is an unmitigated epidemic, and not an absence of these harms. The socioeconomic situation of informal settlements means that respondents may face greater economic precarity than residents of formal urban areas. Even within this sample, the poorest quintile of respondents reported 1.5 times as many contacts as the richest, suggesting an inequitable impact of COVID-19 transmission. This inequity would be exacerbated if socially patterned financial and access barriers inhibit the poor from seeking care for COVID-19[26, 27]. Stringent control measures which cause economic and food insecurity are not likely to be sustainable in the long term if not accompanied by social protection mechanisms.

This study has a number of limitations. In the absence of baseline contact data (i.e. before control measures were put in place), we use empirical matrices from a different area of Kenya and synthetic matrices based on adjusting contact surveys from higher income countries to household and other characteristics in Kenya. Although we adjust these datasets by the age structure of the Kenyan population, other factors such as household size were not reported and may influence number of contacts and therefore pathogen transmission. The pre-COVID-19 setting of Kiti et al. is very different to this sample, not least as estimates place population density around 24 times greater in informal settlements (Kibera: 55000 persons/km^2^) compared to urban Kilifi (2325 persons/km^2^)[28]. Because we would expect contacts to be greater in more densely populated areas, the true reduction in contacts may be more than we estimate here. Although we have a range of background data on respondents from using existing sampling frames, households in the AGI-K and NITISU cohorts were initially selected as having an adolescent residing there in 2015 and 2018, respectively.

Other social contact surveys have used a prospective study design, asking respondents to record contacts in a daily diary[29]. Because we asked respondents to recall contacts from the previous day, these data may be subject to recall bias, although it is not clear in which direction this may act. Furthermore, we impute adjusted child contacts using the comparison studies. An alternative approach, such as that taken by Kiti et al., would have been for respondents to record contacts for children in their household – arranging this was not possible during COVID-19 restrictions. To make the contact survey feasible for phone-based data collection, we simplified the tool for respondents who reported more than ten outside household contacts. We are therefore limited to knowing these contacts’ age and whether the contact was physical or non-physical. Contacts reported in this way were a substantial proportion (70%) of the total sample. The main risk of bias from this may stem from respondents rounding up or down to anchor numbers (e.g. units of ten); figure 1 panel E shows a few respondents cluster around 50 and 100 contacts. Overall, the loss of granularity was beneficial to reducing respondent burden.

We do not calculate the net reproduction number, *R*, but because reported case numbers in Kenya are low, the proportion of the population that is no longer susceptible is likely minimal. We assume that direct contacts are a proxy for effective contacts and therefore transmission, and that transmissibility does not vary by age. In addition, we do not account for the very high proportion of respondents who report that they or their direct contacts wore face masks. Considering these factors would mean *R* is below the *R_0_* estimated here.

## Conclusion

Kenya has implemented strict control measures in response to the COVID-19 pandemic. This study highlights the difficult decisions policymakers face as we find that control measures are likely to have substantially reduced COVID-19 transmission, but also negatively impacted food and economic security of informal settlement residents. This is the first study to measure social contact patterns after COVID-19 control measures have been implemented in sub-Saharan Africa. There is evidence that impacts are inequitable, as the poorest quintile report 1.5 times more contacts than the richest quintile, and 86% of respondents reported complete or partial income losses. Negative and inequitable impacts on economic and food security may mean control measures are not sustainable in the longer term without social protection.

## Data Availability

Data and code fully available at https://github.com/mquaife/kenya_mixing

## Acknowledgments

We are grateful for the excellent research assistance of Roseline Oguta, Hellen Collete Ochola, Emmanuel Mukabi, Carol Olela Adhiambo, Catherine Nduku Mwangi, Wesely Onsongo, Omachi Shawn Ambunya, James Joseph Okwogo, Esther Kariuki, Juliet Nduta Mwangi, Kadija Mohamed Ali, Vidah Achieng Oloo, Lucy Kerubo Nyamwaro. We also acknowledge the contribution of Timothy Abuya, Faith Mbushi, Eva Muluve, James B. Tidwell and Thoai D. Ngo.

## Supplementary file 1: Mixing data collection tool

### MIXING MATRIX

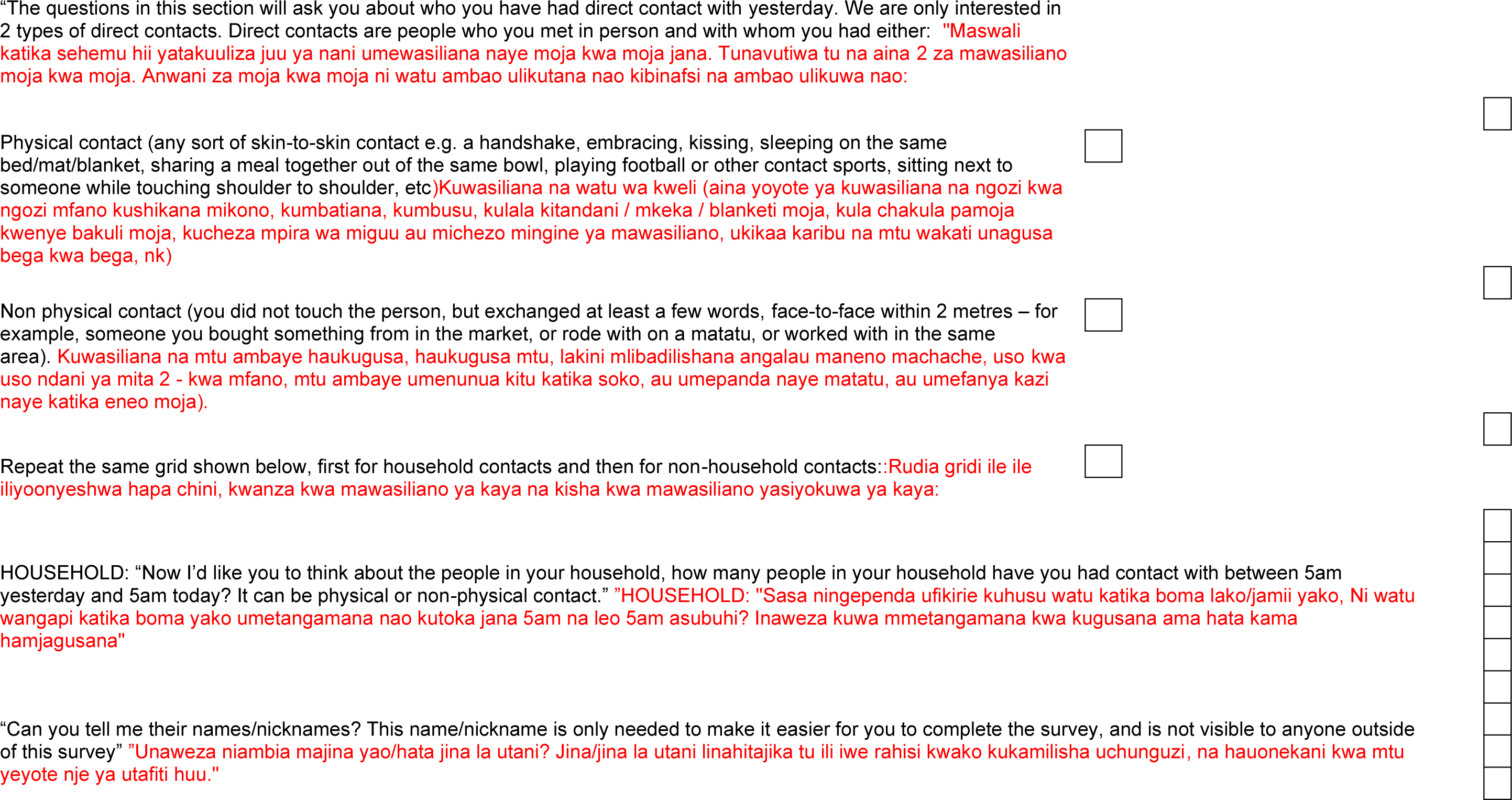
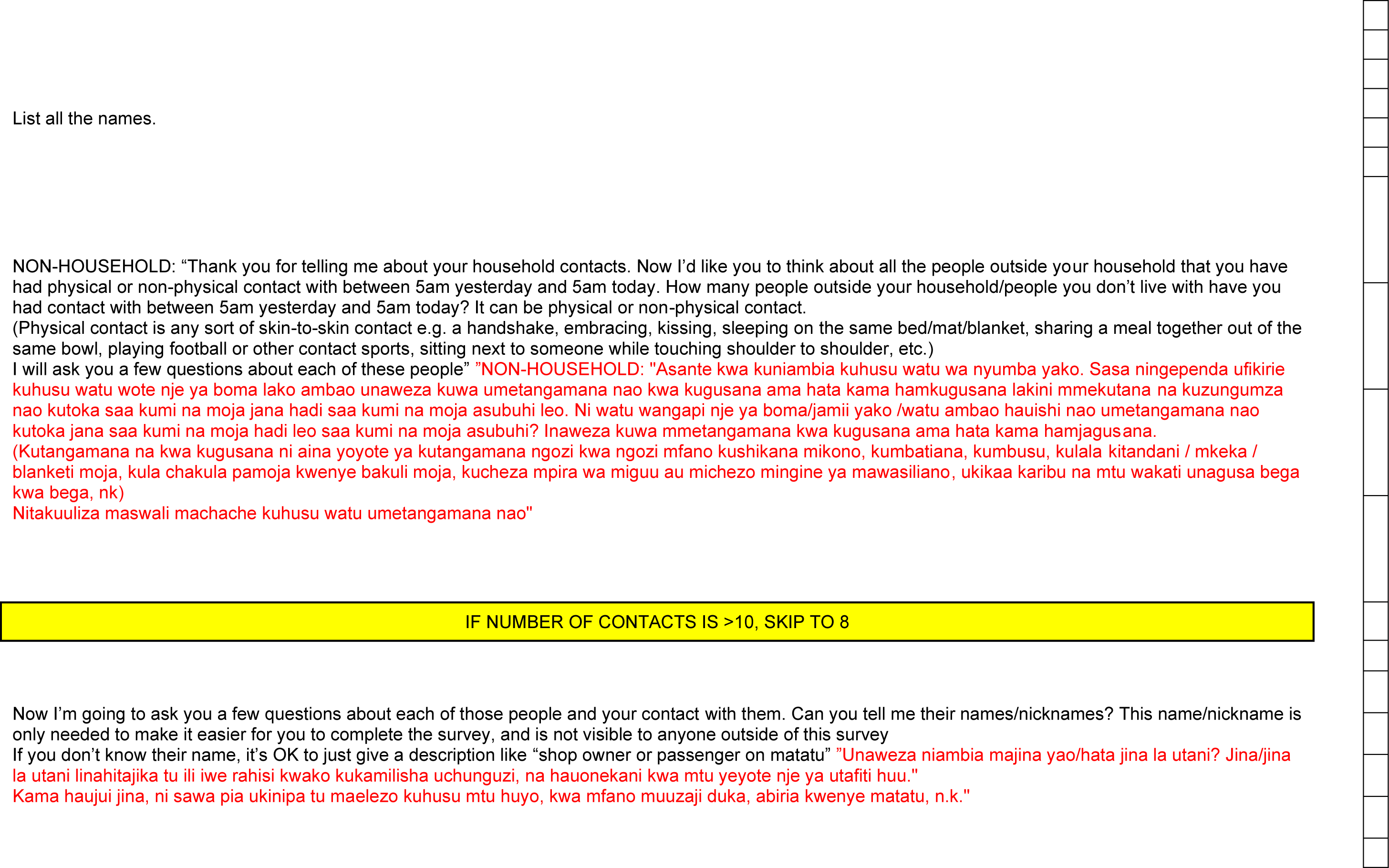

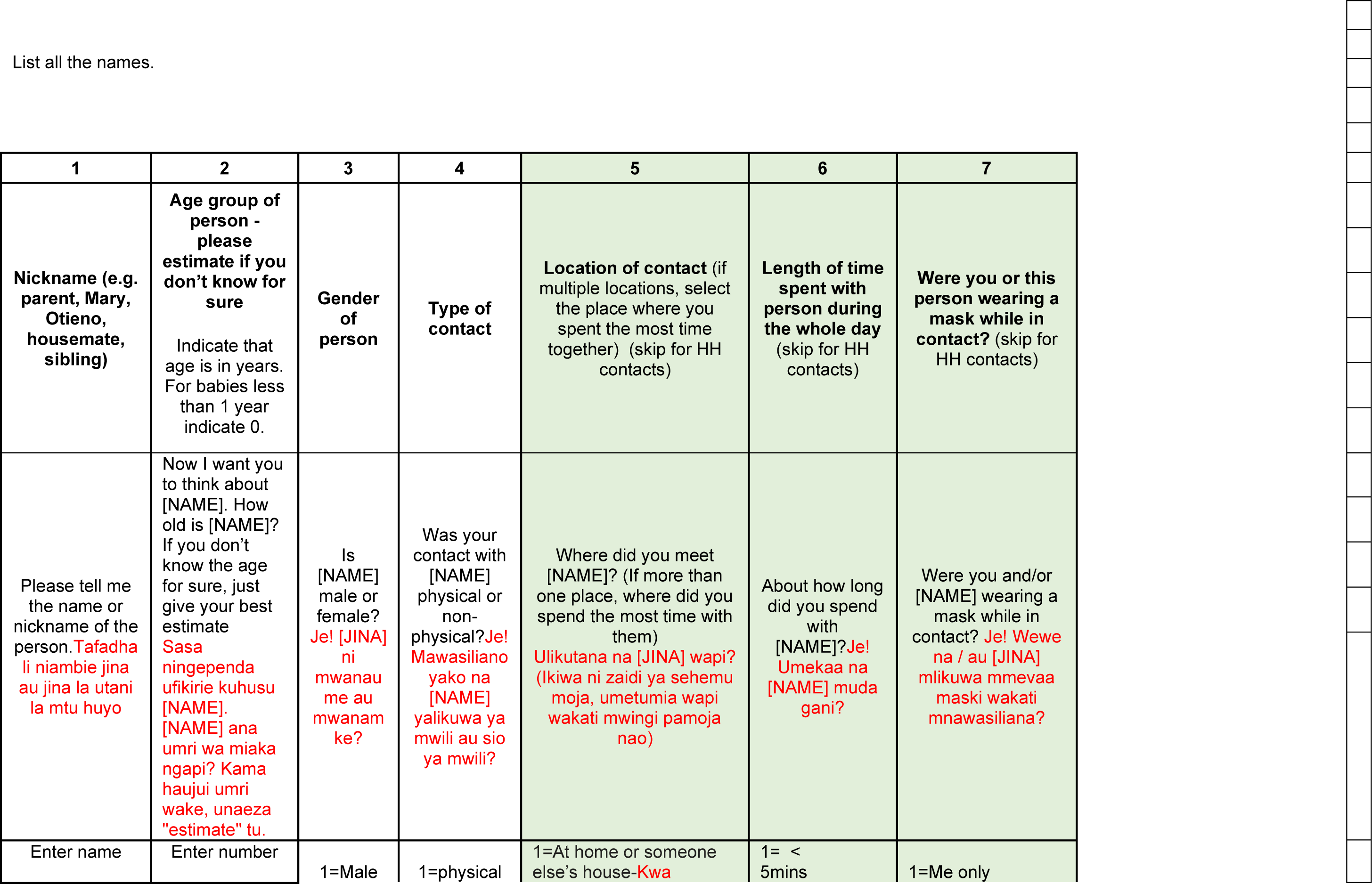
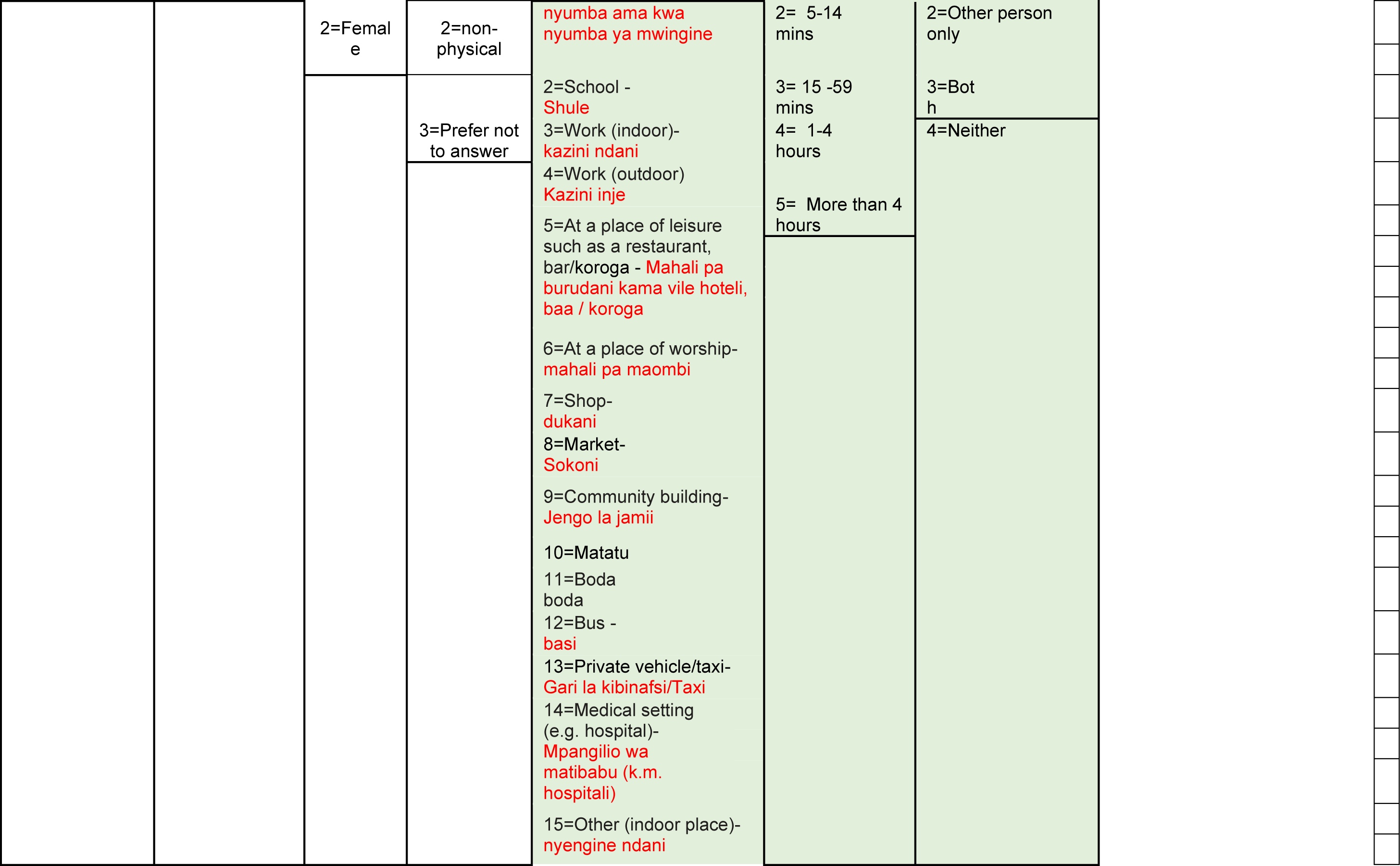
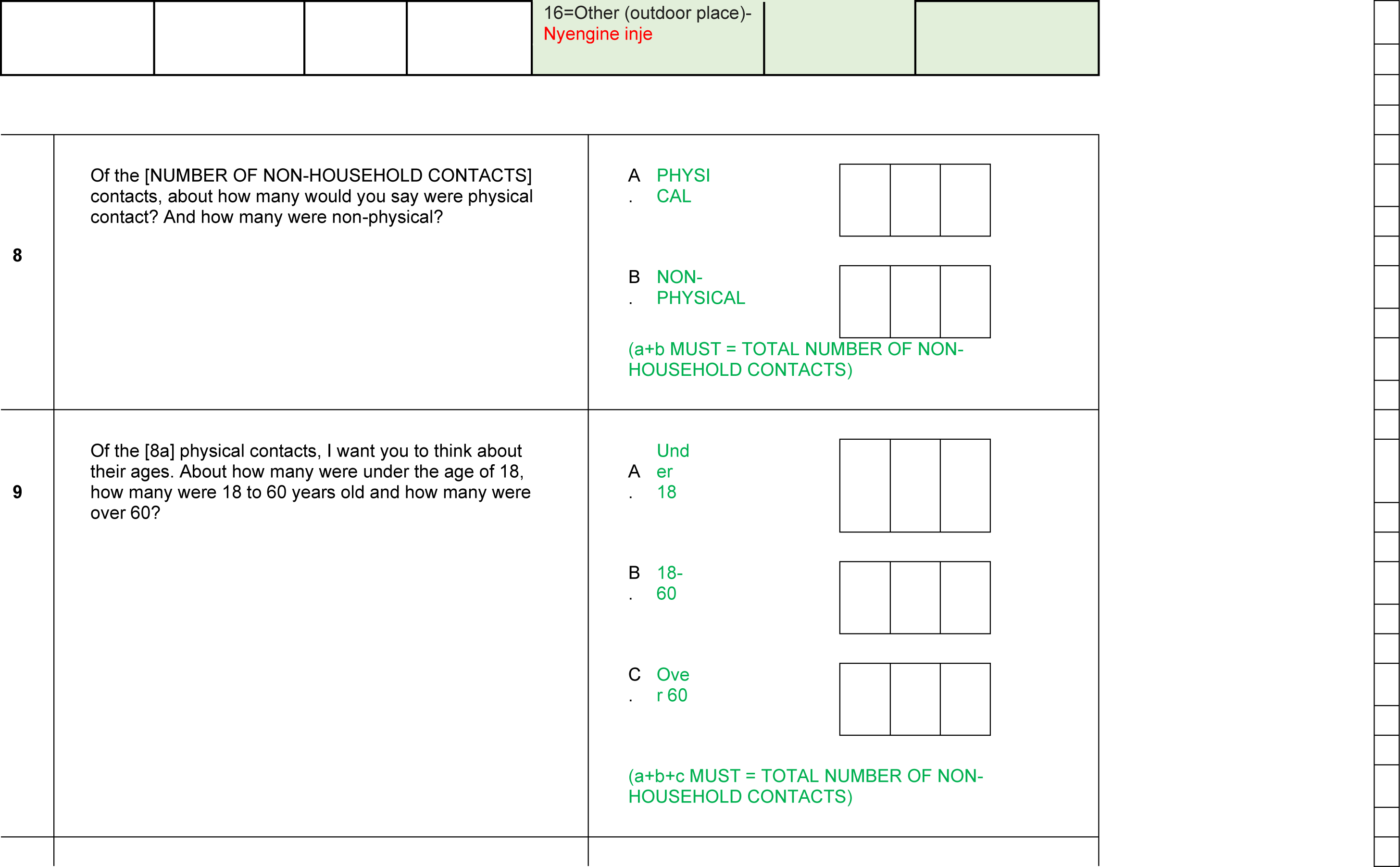
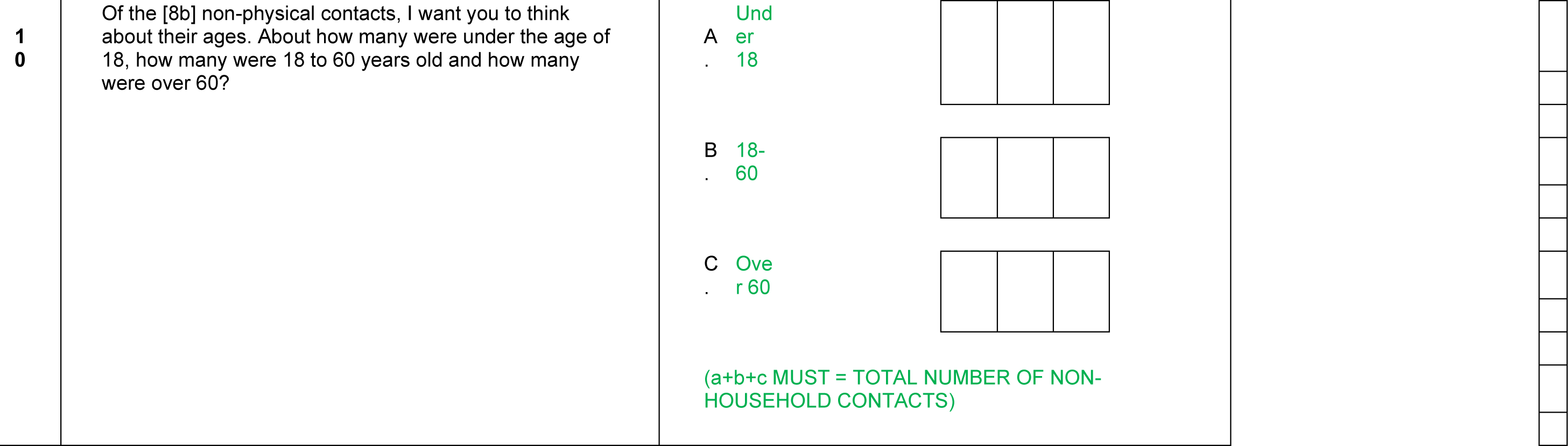

## Supplementary file 2: Measurement of socioeconomic status, and food and economic security

The following characteristics of respondents’ households were used in a principle components analysis to categorise the sample into five wealth quintiles. This categorisation was conducted on the full sample of the of the knowledge, attitudes and perceptions (KAP) survey (n = 1750), of which 213 completed the indepth contacts survey in addition:

1. Piped water supplying dwelling or compound
2. Flushing toilet, to sewer, tank, pit latrine, or elsewhere
3. Toilet not shared with other households
4. Cooking fuel of kerosene, natural gas/LPG, or biogas
5. Finished floor (parquet/polished floor, vinyl/asphalt strips, ceramic tiles, cement, or carpet)
6. Concrete or tiles used in roof
7. Dwelling of more than one room
8. Electricity supply to dwelling or compound
9. Television in household
10. Mobile phone in household

Table S2:1 shows the distribution of respondents by socioeconomic quintile. The distribution of respondents across SES quintiles was similar in the KAP survey and the subset of respondents who were randomly chosen to complete the contacts survey.

**Table S2:1.** Breakdown of socioeconomic quintiles in samples

| Quintile | Respondents<br>(contacts survey) | % | Respondents<br>(KAP survey) | % |
| --- | --- | --- | --- | --- |
| 1 (=poorest) | 36 | 17 | 362 | 21 |
| 2 | 47 | 22 | 343 | 20 |
| 3 | 40 | 19 | 351 | 20 |
| 4 | 71 | 33 | 553 | 32 |
| 5(=richest) | 19 | 9 | 141 | 8 |
| **Total** | **213** |  | **1750** |  |

### Impact on food and economic security

Participants were asked the following questions to elicit the implications of COVID-19 and control measures on food and economic security:

*I want to ask a few more questions about how the Coronavirus pandemic, and the responses of the government and others to try prevent the spread of Coronavirus,’ may have affected you. Your responses will not have an effect on anything you may receive, so please answer as honestly as possible. In the past two weeks, have you experienced any of the following as compared to before the Coronavirus started?*

- *See my family less*
- *See my friends less*
- *Avoid public transport*
- *Complete loss of job/income*
- *Partial loss of job/income*
- *Increased expenses for the household*
- *More housework (cooking, cleaning, caring for children/sick)*
- *More tensions in the household*
- *Increase of crime in your neighborhood?*
- *Experienced more violence outside the house?*
- *Experienced more violence inside the household?*
- *Not purchasing sanitary pads (women only)*
- *Not accessing health care/services/medicines that you would have otherwise needed*
- *Increase in food prices*
- *Other*

## Supplementary file 3: Age adjustment

The mixing matrices of Kiti et al. (2014) [21] and Prem et al. (2017) [19] and (2020) [20] were adjusted for the informal settlement age-distribution. We obtained Kenyan country-level age distribution data from the UN World Population Prospects for 2020. We obtained sub-county age distributions for Kilifi and informal settlements from 2019 Kenyan census data[28]. Datasets are available in the study github repository.

**Figure S3:1.**
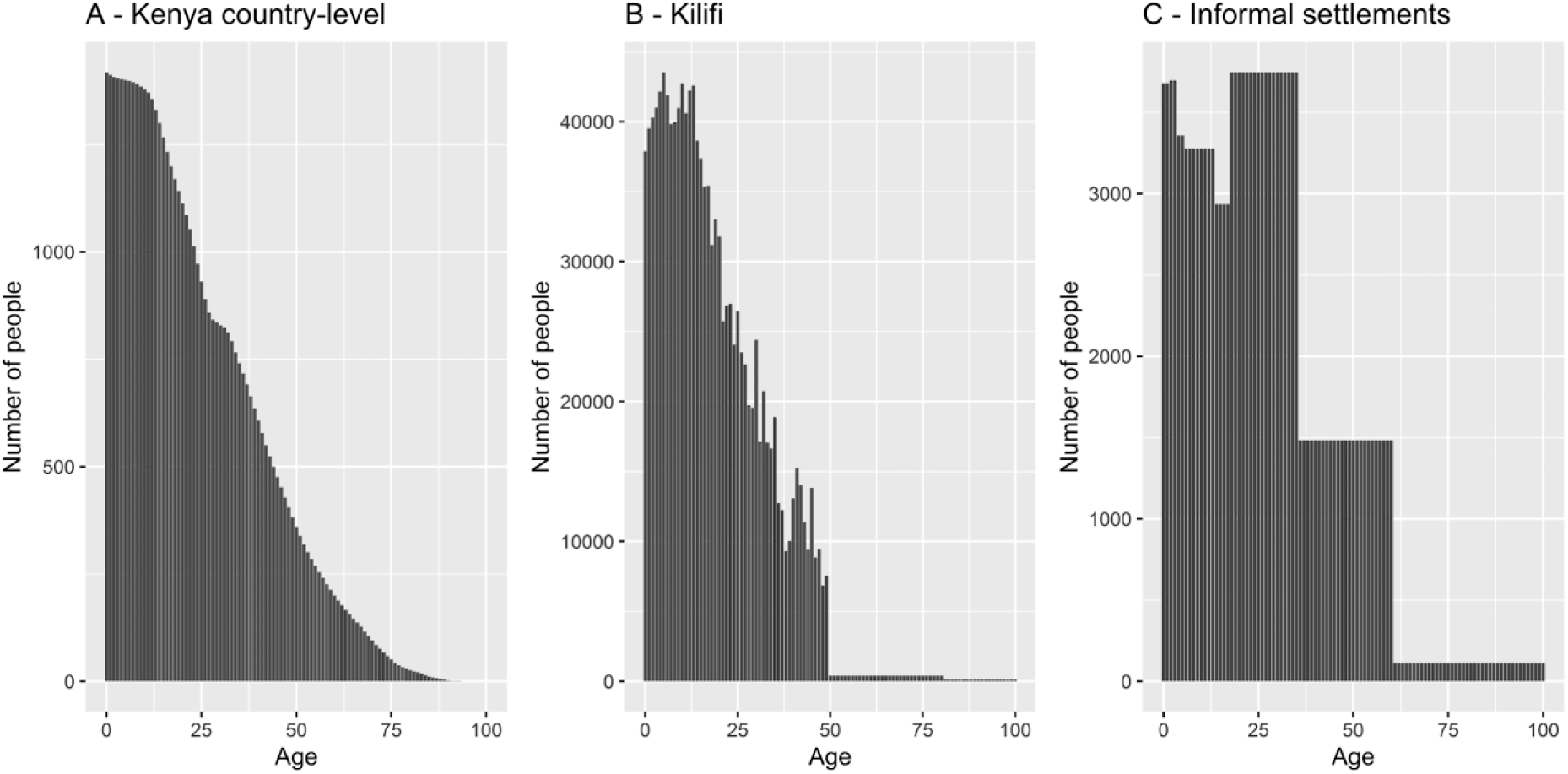
Age distributions for (A) Kenya country-level, (B) Kilifi, and (C) Nairobi informal settlements

## Footnotes

1 Fever, headache, cough, diarrhea, difficulty breathing, loss of taste or smell, tiredness/fatigue, chest pain, chills, rash, dizziness, sneezing, sore throat, myalgia

